# The impact of COVID-19 on the lives and mental health of Australian adolescents

**DOI:** 10.1101/2020.09.07.20190124

**Authors:** Sophie H. Li, Joanne R. Beames, Jill M. Newby, Kate Maston, Helen Christensen, Aliza Werner-Seidler

## Abstract

**Objective:** There has been significant disruption to the lives and mental health of adolescents during the COVID-19 pandemic, but the exact nature of the effects is not known. The purpose of this study was to assess the psychological and lifestyle impact of the pandemic on Australian adolescents, using an online survey, administered during and after the peak of the outbreak (June-July 2020).

**Method:** Self-report surveys were administered online to a sample of 760 Australian adolescents aged 12-18 years old. Surveys assessed worry about contracting COVID-19, behavioral change in response to the pandemic, impact on education, peer and family relationships, lifestyle factors including exercise, technology use and sleep, as well as mental health outcomes including psychological distress, loneliness, health anxiety and wellbeing.

**Results:** Overall, young people expressed significant concern and worry about contracting the virus, and most (>85%) engaged in behaviors to reduce the risk of transmission. Three quarters of the sample reported a worsening of their mental health since the pandemic began, with negative impacts reported by most respondents on learning, friendships and family relationships. More than 40% of young people reported a decrease in exercise and 70% reported an increase in technology use since the outbreak. There were high levels of uncertainty about the future reported by respondents, and their scores on validated measures indicated higher levels of sleep disturbance, psychological distress and health anxiety, and lower levels of wellbeing, relative to normative samples. Reponses on the Kessler Psychological Distress Scale indicated that 48.3% of the sample were experiencing distress consistent with a probable mental illness, which is much higher than pre-pandemic prevalence rates. Effects on mental health were worse among those who reported a previous diagnosis of depression and/or anxiety relative to those without a history of depression and/or anxiety.

**Conclusion:** These results indicate high levels of disruption and psychological distress experienced by adolescents during the current COVID-19 pandemic. Adolescents are already vulnerable to the onset of mental illness at this developmental stage, and the current research underscores the need to find rapid and accessible ways to support adolescent mental health during times of crisis. There is a need for longitudinal research to evaluate the enduring effects of the pandemic on adolescents.

---

As of the 31^st^ of August 2020, there have been more than 24 million COVID-19 cases and 838,924 deaths worldwide^1^, with 25,547 cases and 600 deaths in Australia. As the pandemic and associated restrictions continue, there has been growing concern about the impact on mental health. Studies from around the world have shown that most individuals report increased psychological distress and worsened mental health^2–4^, an effect which seems to be amplified among those with a history of mental illness^5,6^. Although worsened mental health has been documented across the lifespan, several studies have found young adults (aged 18-24 years) are experiencing the greatest deterioration in mental health^3,7^. Very few studies have assessed the impact of the pandemic-related disruption on adolescents^8^, which is concerning because adolescence represents a time of social transformation, marked by an increased need for peer interaction and heightened sensitivity to social stimuli^9^. Adolescents demonstrate increased independence from their families, begin to establish relationships with peers based on shared values and ideas^10^, and become more sensitive to peer acceptance, approval and rejection than either children or adults^11^. Because of this sensitivity, it is likely that social distancing measures, together with school closures, may have a greater negative effect on adolescents. Support for this comes from a recent review of the literature involving more than 50,000 young people that found that social isolation and loneliness significantly increases the risk of mental illness in young people^12^.

Despite the significant potential impact of pandemic-related restrictions on adolescents, little empirical work has addressed this possibility. Four published studies have assessed adolescent mental health (<18 years) in response to the pandemic, three conducted in China and one in the US^13–16^, which collectively show that the prevalence of mental illness (most commonly depression and anxiety) is elevated relative to pre-pandemic prevalence estimates. Evidence of a similar pattern of worsening youth mental health in Australia is emerging. For example, the Kids Helpline (a 24-hour free counselling service for 5-25 year olds), has reported a 40% increase in calls to the service, and the number of young people attending the hospital Emergency Department for self-harm has increased by 33%, both relative to the same period in 2019^17,18^. In addition, young people’s behavior and lifestyles have also been impacted by COVID-19. For example, one study^16^ found that most teenagers reported engaging in at least some social distancing, and another found that 94% of adolescents reported engaging in protective behaviours, such as wearing a mask, hand washing and social distancing^14^. Moreover, worsening sleep and feelings of isolation and loneliness have been reported^19^, as has disruption to learning and education^14^. Together, the available data indicates significant impact on young people’s daily lives.

Beyond these studies, it is unknown how the disruption to daily life has impacted adolescents’ behavior and worries as they relate to COVID-19. The impact on their peer relationships, family relationships, feelings of loneliness, learning and education, lifestyle factors (e.g., exercise, sleep, technology use), and how these relate to mental health, requires investigation to understand the support adolescents require. The overall objective of the current study is to address this gap and answer calls from the scientific community to assess how young people’s lives and mental health has been impacted by the pandemic^20,21^. The first aim of this study was to investigate the psychological impact of the pandemic on adolescents. We included measures of psychological distress, loneliness, health anxiety and wellbeing, which we expected to have worsened, as informed by the emerging literature^13,14,16^. Based on previous studies from the adult literature^5,6^, we expected that those who had a pre-existing history of depression and/or anxiety would show a worse psychological response to the pandemic.

The second aim was to understand how the pandemic and associated containment measures impacted the lives of Australian adolescents. To this end, we collected data on young people’s demographic characteristics, worry about contracting COVID-19, changes to behavior, sleep disturbance, exercise, and technology use. Consistent with past studies, we expected young people to express significant concerns about contracting COVID-19, and to report changes in behavior, sleep patterns, exercise and technology use. A final exploratory aim was to examine the relationship between levels of worry about pandemic, behaviour change, uncertainty about the future, exercise, technology use, sleep, loneliness, wellbeing, psychological distress and health anxiety.

Data was collected from an online survey distributed via social media advertisements. While this method may not produce a representative sample of the entire population^22^, it nonetheless allowed timely access to a significant number of young people while containment measures remained in place. The recruitment period (end of June-beginning of August 2020) included the relaxing of lockdown restrictions across Australia (end of June-beginning of July), with the exception of Melbourne and the state of Victoria, which went into a second lockdown on the 8^th^ of July (Melbourne) and 2^nd^ of August (Victoria).

## Method

### Participants

Participants were recruited via social media and were aged 12-18 years, living in Australia. Data was collected via the Qualtrics platform from 22 June 2020-5 August 2020.

### Ethics approval and consent

The study was approved by the UNSW Human Research Ethics Committee (HC200334). All respondents were required to pass a Gillick Competency Task to ensure the understood the study, and had the capacity to provide *informed* consent, before providing consent^23^.

### Measures

#### Demographics, general health and mental health history

Information was collected on participants’ age, school grade, gender, country of birth, language spoken at home, Aboriginal and Torres Strait Islander status, state of residence and who they lived with. Participants indicated if they had a parent or carer whose job had been impacted by the pandemic. Participants rated their self-rated health^24^, and were asked whether they had a chronic illness, had ever been diagnosed with depression or anxiety by a professional, and current mental health treatments.

#### COVID-19 Exposure, Perceived Risk and Behaviour Change

The items below were generated from a previous survey^25^.

##### COVID-19 Exposure

Participants were asked five yes/no questions about COVID-19 – whether they had been tested/diagnosed, whether a family member or close contact had been diagnosed, and whether they had been required to quarantine for 14 days.

##### Perceived Risk

Participants were asked four questions relating to their perceptions of risk and their concerns about contracting COVID-19. The first question assessed how worried they were about catching COVID-19 on a 5-point scale (not at all – extremely concerned). They then rated perceived likelihood would catch the virus on a visual analogue scale (VAS) from 0 (not at all likely) to 100 (extremely likely), and perceived behavioural control (i.e., how much they thought they could do to protect themselves from catching the virus), on a 0 (I can’t do anything) to 100 (I can do a lot) VAS. The final questions assessed perceived illness severity if they did catch COVID-19 (response options were: no symptoms, mild symptoms, moderate symptoms, severe symptoms, severe symptoms requiring hospitalisation, and severe symptoms leading to death).

##### Behaviour Change

Participants indicated whether they had engaged in eight social distancing and hygiene behaviours over the past week (hand washing, hand sanitising, facemask wearing, avoidance of: going to the shops/touching surfaces/spending time with people from outside the household/going to school and whether they stayed home as much as possible). Response options were on a 5-point scale (1 = not at all to 5 = to all of the time).

#### COVID-19 Impact on Physical and Mental Health, School/Education and Relationships

##### Physical and mental health

Young people were asked, in two separate questions, whether their physical and mental health had been impacted by the pandemic (a lot better, a little better, stayed the same, a little worse, a lot worse).

##### School and education

Respondents indicated whether their school shut down during the pandemic and whether they participated in online learning (both yes/no). Those engaged in online learning reported up to three challenging aspects of completing school work at home, from a forced choice list (not enough family support, parents working and didn’t have time to help, access to technology, speed of internet, harder to learn online, too many distractions, not enough support from teachers, not motivated, other). Participants indicated how much they felt the pandemic had impacted their learning overall (positively, not at all, negatively).

##### Peer relationships

Two questions assessed how the pandemic had impacted participants’ friendships; first, how socially connected they felt to their friends (more connected, no change, less connected); and second, impact on friendships overall (positively, negatively, not at all).

##### Family functioning

Two items assessed young people’s family relationships, in terms of overall impact on relationships with family members (improved, no change, worsened), and impact on family stress levels (less stress, no stress, more stress).

#### Lifestyle Factors

##### Exercise

Participants indicated the number of days they exercised for at least 30 minutes over the previous week (0, 1-2 days, 2-4 days, 5-6 days, every day, don’t know), and to report whether, they exercised more, less, or the same amount as they usually would.

##### Technology use

For the previous week, participants estimated total time spent in front of screens each day, not including time spent on school work (< 1 hour, 1-2 hours, 2-4 hours, 6-8 hours, and > 8 hours). They also reported how much time was used to connect with friends or family (same response options) and whether their use of technology had changed since the pandemic (less than before, the same, more than before).

##### Sleep

Sleep was measured using the Insomnia Severity Index (ISI), which is a seven-item self-report measure of insomnia symptoms over the previous two weeks^26^. Responses are reported on a Likert scale from 0-4 with the following cut-off scores: 0-7 no clinically significant insomnia, 8-14 subthreshold insomnia, 15-21 moderate severity insomnia, 22-28 severe insomnia^26^. The ISI has been widely administered to, and validated in, general adolescent samples^27^.

##### Loneliness

A single item was selected to assess loneliness and was taken from the UCLA Loneliness Scale^28^. Participants were asked to indicate how often they felt alone over the past two weeks (hardly ever, some of the time, often).

##### Uncertainty about the future

A single item was selected to ask participants about their feelings of uncertainty about the future, ranging from ‘not at all’ to ‘extremely’ on a five-point scale where a higher score represents greater levels of uncertainty.

#### Mental Health & Wellbeing

The Kessler-6 (K6) assessed general psychological distress over the past 30 days^29,30^, and the 7-item Warwick Edinburgh Mental Wellbeing Scale – short form SWEMWS;^31^ assessed mental wellbeing over the past two weeks. Both scales have been validated for use in young people^32,33^. The three item Body Preoccupation Scale of the Illness Attitude Scales^34^ was administered to assess health anxiety.

#### Procedure

Participants responded to study advertisements by clicking on a link which took them to the survey landing page. They read the electronic information sheet and consent form, completed the Gillick Competency Task, before accessing the survey. Upon completion, participants were placed in a draw to win one of five $50 vouchers.

#### Statistical Analyses

Demographic and clinical characteristics were reported using descriptive analyses. Where possible, outcomes from standardised measures were compared to normative, general population data. Independent samples *t*-tests compared participants with and without a prior diagnosis of anxiety and/or depression on outcome variables (e.g., K6). Zero order correlations were conducted between worry about COVID, behaviour change, lifestyle and mental health variables.

### Results

In total, 1743 young people viewed the study page, and 945 participants provided consent and started the survey. Of those, 185 did not provide enough data (<90% complete) to be included. The final sample comprised 760 participants.

#### Demographics

Table 1 summarizes participant characteristics. Participants were 14.8 years on average, and ranged in school year from Years 7-12. Most were female (72%), spoke English at home (87.7%) and were born in Australia (88.1%). Participants lived across all states and territories. Almost two-thirds of the sample (63.2%) reported living with two parents, and 50% indicated that their parent or carer’s job had been impacted by the pandemic.

**Table 1.**
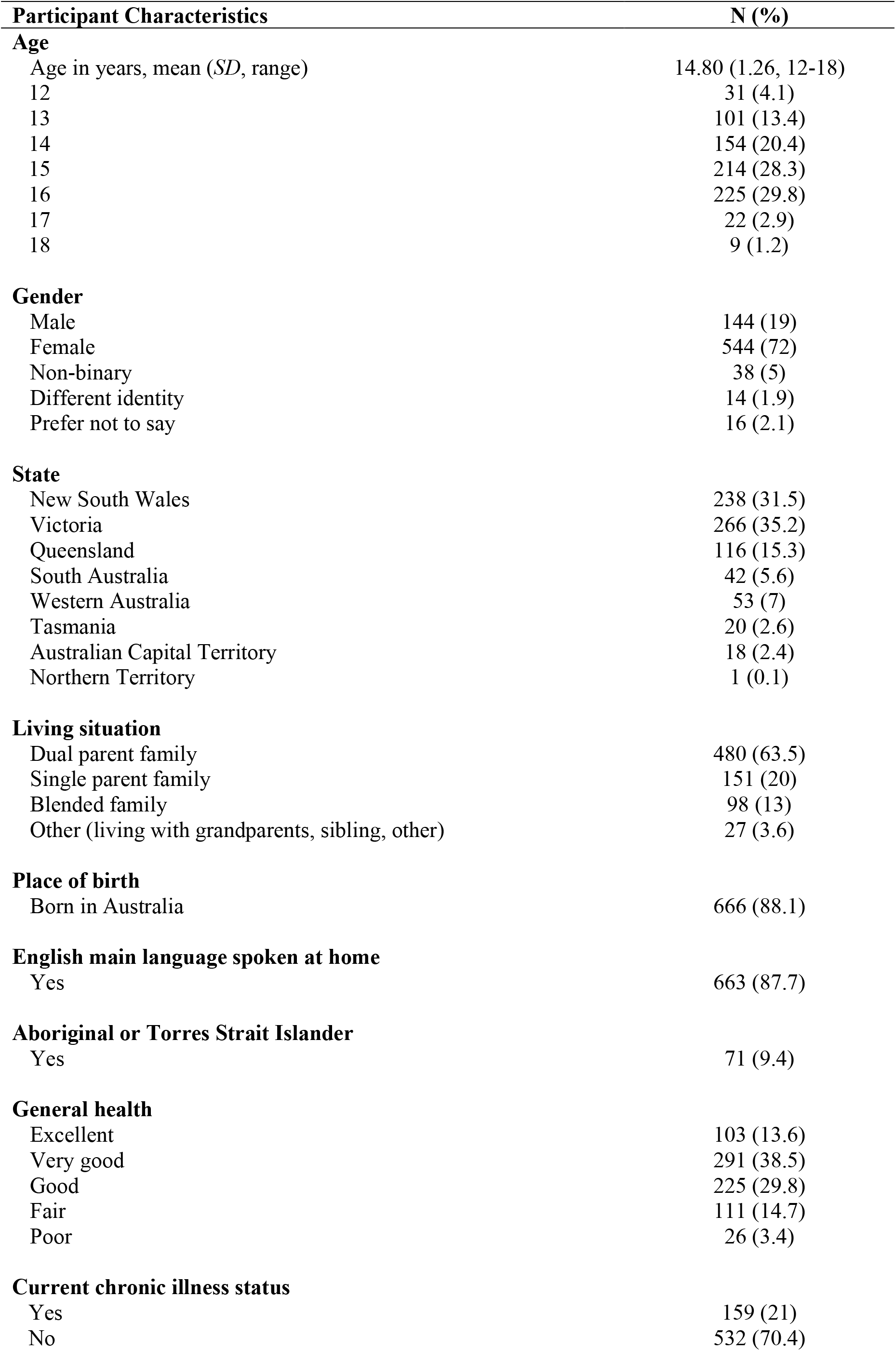
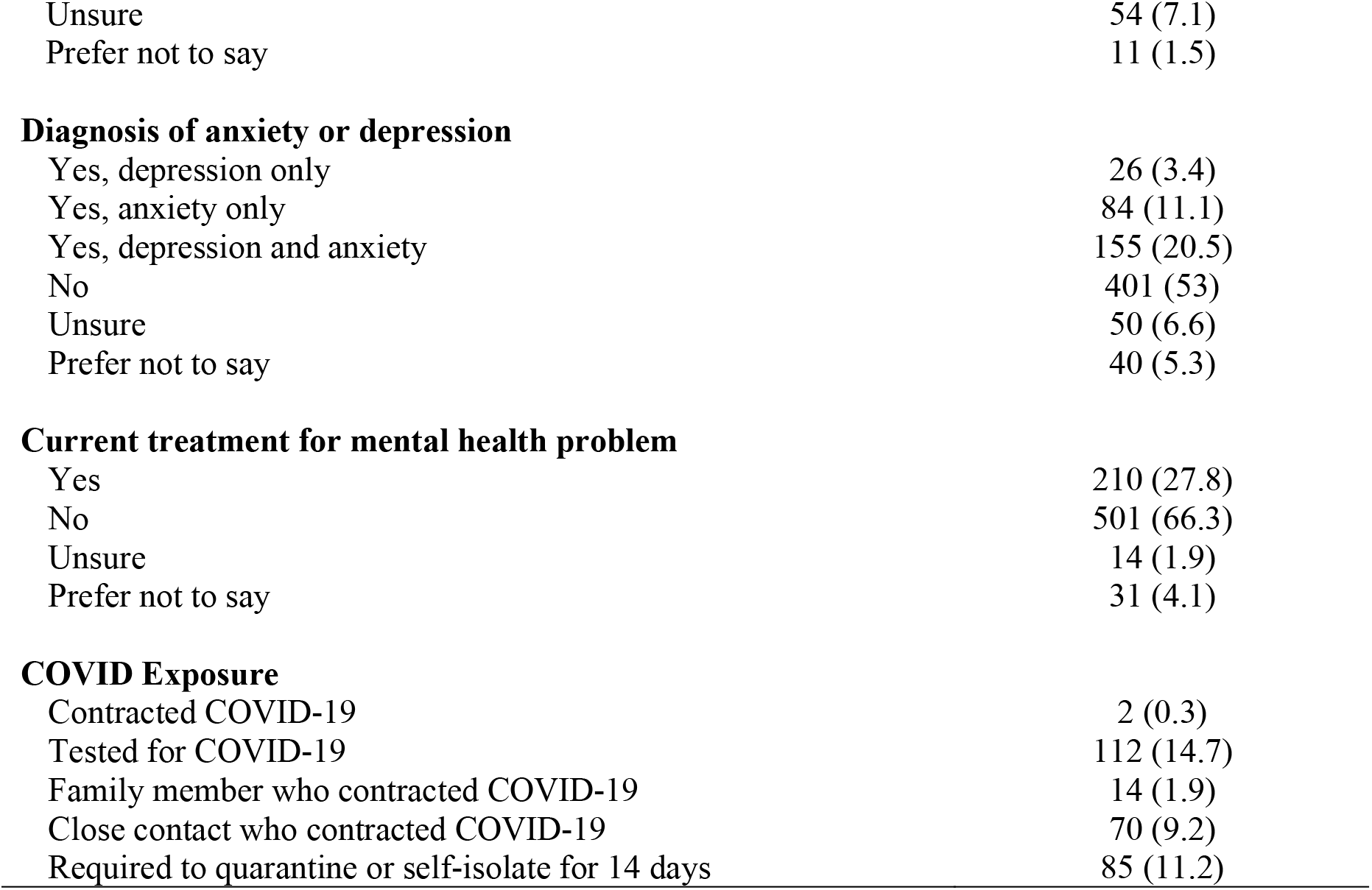
Participant characteristics, clinical history and COVID-19 exposure

#### General Health and Mental Health History

The mean rating for overall health was 3.44/5 (*SD* = 1.01), with most rating their health as either ‘good’ (29.8%) or ‘very good’ (38.5%). Twenty one percent reported a current chronic illness, 34% reported a previous diagnosis of either depression or anxiety, and 27.8% were receiving current mental health treatment (See Table 1).

#### COVID Exposure, Perceived Risk and Behavior Change

##### Exposure

Only 0.3% had a diagnosis of COVID-19. Few participants had a family member who had contracted the virus (1.9%), while just under 10% had a close contact who had the virus.

##### Perceived risk

On average, young people expressed moderate levels of worry about contracting the virus (M = 2.27, *SD* = 0.97), and most (68.5%) were ‘a little concerned’ or ‘ moderately concerned’ about catching the virus. Respondents thought it was relatively unlikely that they would contract COVID-19 (M = 25.29; *SD* = 19.81; scale 0-100), and reported a sense of agency that their behaviors could prevent them contracting the infection (M = 60.75; *SD* = 24.29; scale 0-100). Young people expected if they contracted the virus they would experience either no (5.3%) or mild (39.8%) symptoms, with 37.2% expecting moderate symptoms.

##### Behavior change

Most engaged in protective health-related behaviours over the previous week. Reports of handwashing were high, with 68.6% indicating they washed their hands thoroughly ‘all of the time’ or ‘most of the time’ as was sanitizer use (72.1% all or most of the time). Most (85.6%) respondents avoided touching objects or surfaces touched by other people to at least some extent, and 30% reported wearing a facemask. Respondents also avoided places to reduce the spread, with 82.1% avoiding the shops at least some of the time (43.3% reported avoiding shops all or most of the time), 81.3% avoiding leaving the house (48.4% always or most of the time), and 70.4% avoiding spending time with people outside their household (35.1% always or most of the time). Half reported not going to school in the previous week due to the pandemic.

#### COVID-19 Impact

##### Physical and mental health

See Figure 1. More than half of the participants indicated that their physical health had worsened during the pandemic. Approximately a third reported no change, and few reported an improvement. The impact of the pandemic on mental health revealed different results; most (75%) young people reported a negative effect on their mental health. Few reported no change or an improvement.

**Figure 1:**
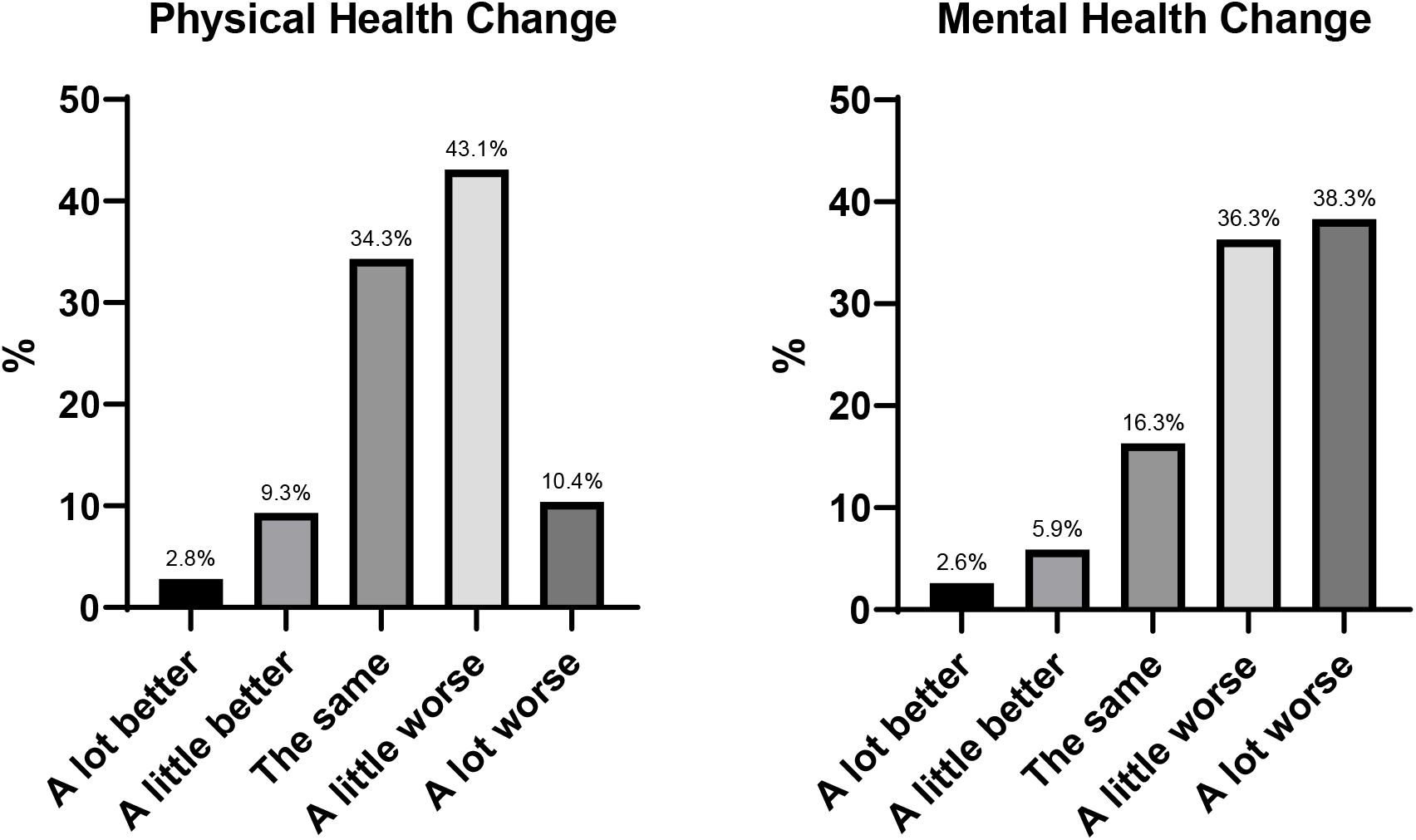
Physical and mental health change since the pandemic began

##### School and education

Most (87%) reported that their schools closed during the pandemic, and the majority (95.1%) engaged in online learning instead. The top three challenges to online learning were: i) a lack of motivation; (ii) too many distractions at home, and iii) it being more difficult to learn via online platform relative to face-to-face. A lack of support from schoolteachers, increased school workload, feelings of loneliness and slow internet was also noted. Overall, two in three young people (62.6%) felt that the pandemic had negatively impacted their learning, with 22% indicating no change and 14.9% reporting a positive impact.

##### Peer relationships

See Figure 2. Most respondents reported feeling less connected to their friends. There seemed to be a degree of stability to adolescents’ friendships, with about half of the sample reporting no overall impact on their friendships.

**Figure 2.**
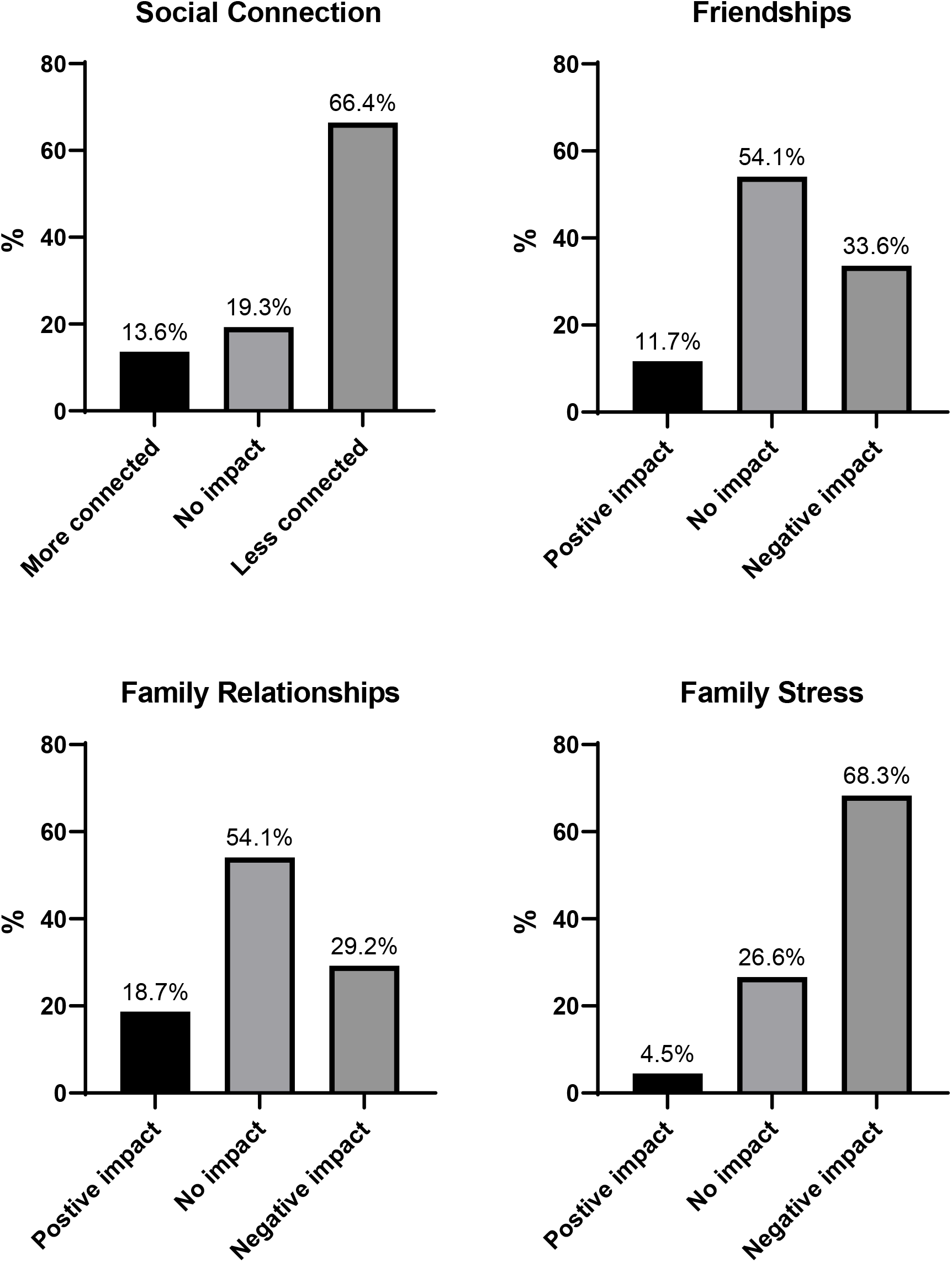
Impact of pandemic on peer and family relationships

##### Family functioning

See Figure 2. Approximately one third of respondents reported a worsening of family relationships, and most young people reported a worsening of family stress.

#### Lifestyle Factors

See Table 2.

**Table 2.** Frequency and descriptive data for lifestyle and mental health outcomes

| **Lifestyle and Mental Health Outcomes** |  |  |
| --- | --- | --- |
| Exercise (# ≥30 mins exercise sessions in past week) | **N (%)** |  |
| None | 154 | (20.3) |
| 1-2 days | 236 | (31.1) |
| 3-4 days | 169 | (22.2) |
| 5-6 days | 102 | (13.4) |
| 7+ days | 76 | (10) |
| Exercise change due to the pandemic |  |  |
| Less than usual | 308 | (40.5) |
| Same as usual | 326 | (42.9) |
| More than usual | 121 | (15.9) |
| Technology Use<br>(time spent on screens each day for non-school purposes) |  |  |
| <1 hour | 11 | (1.4) |
| 1-2 hours | 33 | (4.3) |
| 2-4 hours | 178 | (23.4) |
| 4-6 hours | 241 | (31.7) |
| 6-8 hours | 151 | (19.9) |
| >8 hours | 141 | (18.6) |
| Technology Use – Social Connection<br>(amount of time spent connecting with friends or family) |  |  |
| <1 hour | 173 | (22.8) |
| 1-2 hours | 243 | (32) |
| 2-4 hours | 196 | (25.8) |
| 4-6 hours | 96 | (12.6) |
| 6-8 hours | 23 | (3) |
| >8 hours | 24 | (3.2) |
| Technology use change due to the pandemic |  |  |
| Less than usual | 81 | (10.7) |
| Same as usual | 128 | (16.8) |
| More than usual | 547 | (72) |
| Loneliness (frequency of feeling alone) |  |  |
| Hardly ever | 130 | (17.1) |
| Some of the time | 233 | (30.7) |
| Often | 391 | (51.4) |
| Uncertainty about the future |  |  |
| Not at all | 54 | (7.1) |
| A little uncertain | 218 | (28.7) |
| Moderately uncertain | 227 | (29.9) |
| Very uncertain | 158 | (20.8) |
| Extremely uncertain | 99 | (13) |
| **Standardised Questionnaires** | **M (SD)** | **Range** |
| Insomnia Severity Index | 12.09 (6.03) | 0-28 |
| Short Warwick-Edinburgh Mental Wellbeing Scale | 18.97 (5.87) | 7-35 |
| Kessler-6 Scale (Psychological Distress) | 18.08 (6.63) | 6-30 |
| Illness Preoccupation Scale (Health Anxiety) | 4.72 (4.72) | 0-12 |

##### Exercise

Twenty percent reported zero instances of at least 30 minutes exercise during the previous week. Approximately half of respondents indicated that they exercised for 30 minutes on at least 1-2 days. Most young people reported either a decrease in exercise since the pandemic began, or no change.

##### Technology Use

Most young people reported either 2-4 hours or 4-6 hours daily screen use. There was a still a significant proportion of young people reporting higher levels of use (>8 hours) each day, with 72% reporting increased technology use to connect with others, usually spending around 4 hours online for interaction. Almost three quarters of the sample reported increased technology use since the start of the pandemic.

##### Sleep disturbance

The mean score on the ISI was 12.09 (*SD* = 6.03). Just under half of the sample (40.5%) reported subthreshold insomnia, while more than a quarter (28.7%) reported moderate severity insomnia and a further 6.4% reporting severe insomnia.

##### Loneliness

Just over half (51.4%) of the sample reported frequently experiencing feelings of loneliness. A further third reported feeling alone some of the time..

##### Uncertainty about the future

Ninety-three percent of respondents reported some degree of uncertainty about the future, with approximately one third (33.8%) reporting very or extreme levels of uncertainty.

#### Mental Health and Wellbeing

##### Mental health

The mean score on the K6 was 18.08 (*SD* = 6.63; Range 6-30), with just under half of the sample (48.3%) scoring above the threshold that indicates psychological distress indicative of mental illness.

##### Wellbeing

Mean wellbeing score was 18.79 (*SD* = 5.87; Range 7-35), with a higher score indicating greater levels of wellbeing.

##### Health anxiety

On the screening survey for health anxiety, the Body Preoccupation Scale of the Illness Attitude Scales, 40.1% of young people scored above the clinical cutoff which indicates severe health anxiety.

#### Comparison between adolescents with and without a prior mental health diagnosis

Young people with and without a self-reported diagnosis of depression and/or anxiety were compared on outcomes (See Table 3). After removing those who did not know if they had a diagnosis or chose not to say, 60% percent of the remaining sample reported no history of mental illness, and 40% reported a previous diagnosis of depression or anxiety (3.9% depression only, 12.6% anxiety only, and 23.3% both depression and anxiety). These categories were collapsed for the subsequent analyses into presence or absence of a diagnosed depression and/or anxiety. Participants with a history of depression or anxiety reported worse mental and physical health on all measures (see Table 3), and reported lower levels of exercise, great use of technology, poorer sleep, higher levels of loneliness, uncertainty about the future, psychological distress, health anxiety and lower levels of wellbeing (all *p*s <0.05; see Table 3 for full statistics).

**Table 3.**
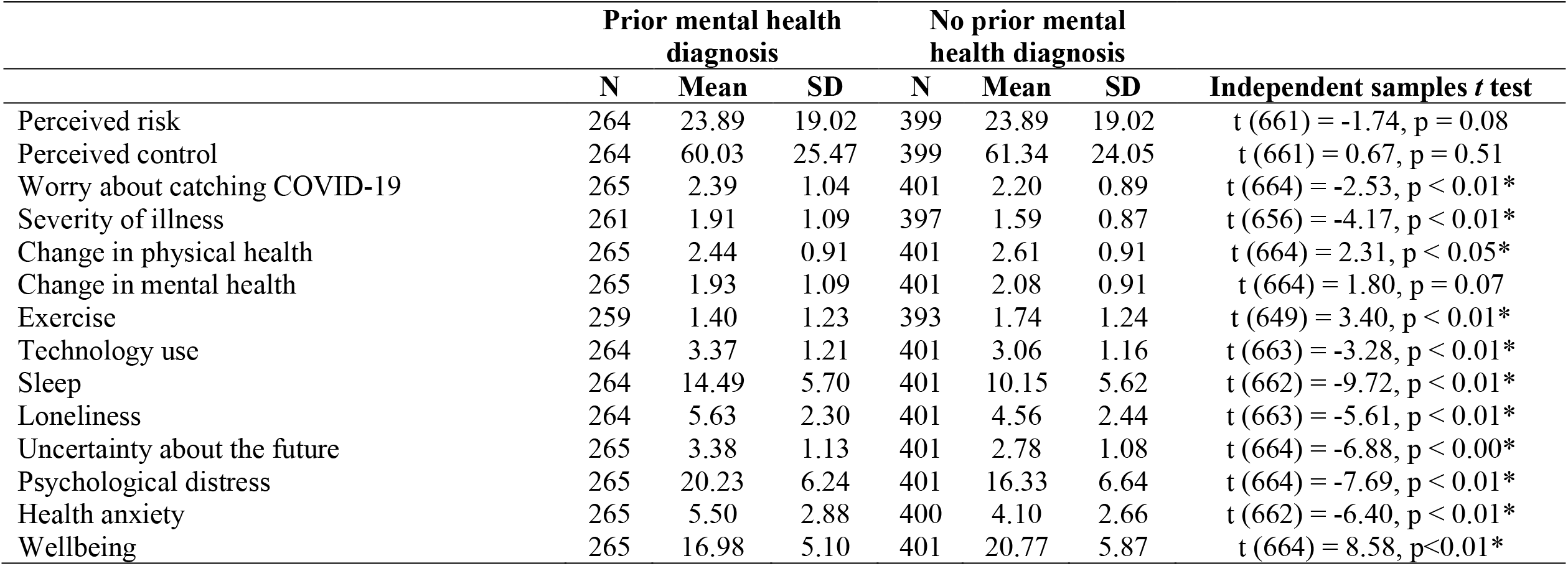
Comparison between respondents with and without a self-reported mental health diagnosis of depression and/or anxiety

|  | Prior mental health diagnosis |  |  | No prior mental health diagnosis |  |  | Independent samples *t* test |
| --- | --- | --- | --- | --- | --- | --- | --- |
|  | N | Mean | SD | N | Mean | SD |  |
| Perceived risk | 264 | 23.89 | 19.02 | 399 | 23.89 | 19.02 | $t(661) = -1.74, p = 0.08$ |
| Perceived control | 264 | 60.03 | 25.47 | 399 | 61.34 | 24.05 | $t(661) = 0.67, p = 0.51$ |
| Worry about catching COVID-19 | 265 | 2.39 | 1.04 | 401 | 2.20 | 0.89 | $t(664) = -2.53, p < 0.01^*$ |
| Severity of illness | 261 | 1.91 | 1.09 | 397 | 1.59 | 0.87 | $t(656) = -4.17, p < 0.01^*$ |
| Change in physical health | 265 | 2.44 | 0.91 | 401 | 2.61 | 0.91 | $t(664) = 2.31, p < 0.05^*$ |
| Change in mental health | 265 | 1.93 | 1.09 | 401 | 2.08 | 0.91 | $t(664) = 1.80, p = 0.07$ |
| Exercise | 259 | 1.40 | 1.23 | 393 | 1.74 | 1.24 | $t(649) = 3.40, p < 0.01^*$ |
| Technology use | 264 | 3.37 | 1.21 | 401 | 3.06 | 1.16 | $t(663) = -3.28, p < 0.01^*$ |
| Sleep | 264 | 14.49 | 5.70 | 401 | 10.15 | 5.62 | $t(662) = -9.72, p < 0.01^*$ |
| Loneliness | 264 | 5.63 | 2.30 | 401 | 4.56 | 2.44 | $t(663) = -5.61, p < 0.01^*$ |
| Uncertainty about the future | 265 | 3.38 | 1.13 | 401 | 2.78 | 1.08 | $t(664) = -6.88, p < 0.00^*$ |
| Psychological distress | 265 | 20.23 | 6.24 | 401 | 16.33 | 6.64 | $t(664) = -7.69, p < 0.01^*$ |
| Health anxiety | 265 | 5.50 | 2.88 | 400 | 4.10 | 2.66 | $t(662) = -6.40, p < 0.01^*$ |
| Wellbeing | 265 | 16.98 | 5.10 | 401 | 20.77 | 5.87 | $t(664) = 8.58, p < 0.01^*$ |

**Supplementary Table:** Correlational Table including COVID variables, lifestyle factors and mental health and wellbeing

|  | **COVID**<br>**Worry** | **Behavior**<br>**Change** | **Uncertainty** | **Exercise** | **Screen**<br>**Time** | **Screen**<br>**Connection** | **Sleep** | **Loneliness** | **Wellbeing** | **Health**<br>**Anxiety** | **Distress** |
| --- | --- | --- | --- | --- | --- | --- | --- | --- | --- | --- | --- |
| ***COVID Variables*** |  |  |  |  |  |  |  |  |  |  |  |
| Worry about contracting COVID | - |  |  |  |  |  |  |  |  |  |  |
| Behavior change | .33** | - |  |  |  |  |  |  |  |  |  |
| ***Lifestyle and other factors*** |  |  |  |  |  |  |  |  |  |  |  |
| Uncertainty about the future | .32** | .13** | - |  |  |  |  |  |  |  |  |
| Exercise | -.02 | -.01 | -.13** | - |  |  |  |  |  |  |  |
| Overall screen time | .01 | .13** | .18** | -.20** | - |  |  |  |  |  |  |
| Screen time to connect with others | .02 | -.01 | -.01 | .06 | .05 | -.05 |  |  |  |  |  |
| Sleep | .11** | .15** | .32** | -.10** | .17** | -.05 | - |  |  |  |  |
| ***Mental Health & Wellbeing*** |  |  |  |  |  |  |  |  |  |  |  |
| Loneliness | .07 | .17** | .28** | -.13** | .11** | -.19** | .31** | - |  |  |  |
| Wellbeing | -.08* | -.07 | -.41** | .21** | -.18** | .14** | -.52** | -.59** | - |  |  |
| Health Anxiety | .23** | .09 | .26** | -.06 | .02 | -.01 | .20** | .20** | -.17** | - |  |
| Psychological Distress | .17** | .15** | .44** | -.12** | .44** | -.04 | .53** | .53** | -.63** | .28* | - |

Note: Zero order correlations presented. \*\* = significant at .01 level; \* = significant at .05 level.

#### Association between COVID-fears, behaviour, lifestyle factors and mental health

See Supplementary materials for results. There was a significant positive correlation between fears about contracting COVID-19, and behaviour change, feelings of uncertainty about the future, poor sleep, psychological distress, and health anxiety (all *p*s<.01). Greater exercise was associated with lower levels of screen time, better sleep, lower levels of psychological distress and a greater sense of wellbeing (*p*s<.01). Overall screen time was associated with more psychological distress; however, when screen time was used to connect with friends and family, the relationship with distress was no longer evident, and increased screen time for connection was associated with lower levels of loneliness and higher levels of wellbeing (*p*s<.01). Finally, as expected, all the mental health variables were significantly associated, with positive correlations between loneliness, health anxiety and psychological distress, and an inverse association between these variables and wellbeing being detected.

## Discussion

This survey of 760 Australian adolescents aged 12-18 years provides the first insight into the disruption experienced following the outbreak of the COVID-19 pandemic. Although respondents had little direct or indirect experience with COVID-19, more than three quarters were worried about contracting the virus, replicating findings with adults^6^ and adolescents^14^. Most young people believed they could reduce their risk of contracting COVID-19 and engaged in behaviors to lower their (e.g., handwashing and social distancing), in contrast to common portrayals by the media suggesting young people are not compliant with restrictions e.g.,^35^. Our results showed that worry about contracting COVID-19 was associated with greater levels of overall behaviour change, demonstrating a practical benefit of some degree of worry.

This study also sheds light on the disruption and impact of the pandemic on young people’s lives. Almost the whole sample (>95%) had engaged in online learning, and most reported a negative impact. At the time our survey was undertaken, schools and families had to adapt to online learning from home with little-to-no preparation time. While meta-analyses have shown that with optimal delivery and support, online formats can be as effective as face-to-face in terms of learning outcomes for adults^36^, there is no evidence to suggest this is the case for young people. Indeed, because school education is primarily face-to-face, data about the effectiveness of online schooling is lacking. Online learning requires a greater level of independence, motivation, and discipline than classroom learning and these are skills which young people may not have fully developed^37^.

Participants reported a negative effect of the pandemic on friendships, and feelings of loneliness, which were associated with higher psychological distress and lower wellbeing. Adolescents are at a crucial stage of development involving the formation of a sense of self and identity through shared interests and values with their peers^38^. Given a lack of social connection has negative consequences on social and cognitive development^9^, and loneliness increases the risk of the development of depression and other disorders^12^, mental health prevention and intervention efforts need to focus on improving social connection, particularly in areas that have containment measures in place for prolonged periods. Until restrictions are lifted, the use of technology to connect with others might mitigate the potential disruption to adolescents’ social needs. With widespread smartphone use^39^, it is reassuring that studies have found that core components of quality face-to-face interactions can be replicated online^40^. Consistent with expectations, our study found that nearly three quarters of participants reported increased use of technology to connect with others and this was associated with lower levels of loneliness and a greater sense of wellbeing. This finding aligns with research from China showing increased smartphone and social media use at the height of the pandemic ^15^. Most young people in our study reported spending about 4 hours a day connecting with others online. Whether this will mitigate potential long-term consequences of social deprivation associated with lockdowns will need to be addressed by future longitudinal studies.

Most respondents indicated that the pandemic had increased stress levels within their family and half reported an impact on the job of a parent or carer. Viewed in this context, together with adjusting to online learning and the requirement for parents to manage their own professional responsibilities with caring and supporting their child’s learning, it is not surprising this has been a stressful time for families. This aligns with findings from around the world that are beginning to highlight higher levels of stress and mental ill-health experienced by certain sub-groups of the population, including women, who are more likely to be in caring roles, as well as parents and young people^3,7^. Despite this increased family stress, > 50% indicated that their relationships with family members had remained unchanged, and an additional 18.7% reported an improvement. It could be the case that in some families, more time at home with family members has some advantages, and feeling more connected to loved ones is a benefit of COVID-19 that has been documented in the adult literature^41^.

Encouragingly, >50% of the young people continued with or increased their regular levels of exercise. There are well-documented links between exercise and reduced risk of depression and anxiety across the lifespan^42^ and the relationship between exercise and lower levels of psychological distress and great wellbeing in our sample is consistent with this broader literature^15^. Given that even the strictest lockdowns in Australia have allowed for up to 1 hour of exercise per day, exercise could be promoted in public health campaigns as a means to prevent deteriorating mental health should the lockdown and period of restrictions continue. Relatedly, a significant proportion of young people also reported increased difficulty sleeping, and exercise is one way in which sleep quality and duration can be improved^43^, which was supported by our correlational analyses.

Of concern was that about half of the sample reported a worsening in their physical health since the pandemic began, while 75% reported a negative effect on their mental health. The worsening of mental health in our sample was markedly consistent to that of a recent adult Australian survey^6^, which found that 78% of their respondents had reported worsened mental health. Overall, young people reported greater psychological distress and lower levels of wellbeing relative to normative data available from population surveys conducted prior to the pandemic^44,45^, with rates of psychological distress indicative of probable mental illness increasing almost two fold from 24.3% before the pandemic^45^, to 48.3% in this survey. This finding accords from the studies from China showing elevated mental illness^8^. It is important to note that the K6 is not a diagnostic interview and so without information about the exact duration of the symptoms, degree of interference in daily life and distress to the individual, drawing diagnostic conclusions is not possible. We also cannot rule out the heightened levels of psychological distress as being at least in part attributable to sampling bias, inherent with online surveys. With these limitations in mind, our data nonetheless suggest that there are significantly elevated rates of psychological distress^45^, providing insight into the acute effects of the COVID-19 pandemic on adolescent mental health.

Over one third (34%) of participants had a self-reported history of depression or anxiety, which is comparable to lifetime prevalence estimates for adolescents^46^. Consistent with the adult literature^5,6^, our results showed that relative to those without a history of depression or anxiety, those with a history experienced heightened levels of loneliness, greater trouble sleeping, more uncertainty about the future, higher levels of health anxiety, greater psychological distress and lower levels of wellbeing among those who had a previously diagnosed mental illness. These finding advance knowledge of youth mental health by showing an exacerbation of mental illness among those with a history of anxiety and/or depression.

The findings reported in this study underscore the need for a proactive mental health response to support young people through this tumultuous and disruptive time in their lives. High levels of psychological distress reported in this study are consistent with increased demands on youth mental health services e.g.,^18^. Readily accessible interventions are needed for young people that are engaging and effective, with a focus on treatment, *and* prevention. Digital mental health interventions, and blended modalities with combine digital with clinician support hold this promise^47^.

There are several study limitations. First, as a convenience sample recruited online due to the ease of rapid administration, 72% were female, limiting the generalizability of findings to the broader Australian adolescent population. The importance of sampling approach has been noted as a key concern during COVID-19^22^. Follow-up studies should use diagnostic assessments, to provide an independent assessment of mental health. The study was cross sectional and so causal conclusions cannot be drawn. We will be continuing to follow this sample longitudinally to learn about how their lives and mental health change over time in response to the pandemic.

## Data Availability

Given the sensitive nature of this data and ethical guidelines, it is not publicly available.

## Acknowledgements

Sincere thanks to all the study participants for taking part in this survey. Thanks also to Iana Wong for assisting with Qualtrics programming, to Dr Samantha Spanos for survey testing and to Dr Lyndsay Brown for providing feedback on the manuscript.

